# Pivotal role of early coronary microvascular dysfunction in Takotsubo Syndrome

**DOI:** 10.1101/2023.08.18.23294289

**Authors:** Gao Jing Ong, Farnaz Jalili, Gnanadevan Mahadavan, John D Horowitz

## Abstract

**Background:** Takotsubo Syndrome (TTS) generally presents like a form of acute coronary syndrome, with variable extents of coronary flow retardation and concomitant release of markers from damaged endothelial glycocalyx (eGC). Whilst systemic hypotension often develops early, there is also rapid emergence and slow resolution of left ventricular (LV) dysfunction. It remains uncertain whether these hypotensive and LV functional complications reflect severity of early coronary vasculitis.

**Methods:** 284 patients admitted to 3 South Australian hospitals from May 2008 to May 2021 with a diagnosis of TTS were evaluated. Coronary flow velocity was measured using the corrected TIMI frame count. Putative correlations between extent of acute coronary flow retardation and that of acute impairment of LV systolic function, as measured on global longitudinal strain (GLS: primary hypothesis), were determined. Other parameters of acute disturbance of homeostasis, including extent of eGC shedding and of myocardial edema, and residual impairment in GLS and quality of life at 3-months’ follow-up, were correlated with extent of flow retardation. We also evaluated correlates of extent of acute systemic hypotension. Results were analysed via univariate followed by multivariate analyses.

**Results:** The patients studied exhibited mild coronary flow retardation relative to previously described norms at early angiography. On univariate analyses, corrected TIMI frame count correlated with extent of acute impairment of GLS (r=0.31, p=0.003) and this association persisted on backwards stepwise multiple logistic regression (β=0.52, p=0.03). Flow retardation also correlated with preservation of renal function (β=0.50, p=0.02), but tended to vary inversely with C-reactive protein (CRP) concentrations (β=-0.44, p=0.06). There were no significant associations between acute TIMI frame count and other acute or 3-months’ parameters. Neither TIMI frame count nor acute GLS predicted minimal systolic blood pressure.

**Conclusions:** The data demonstrated a strong association between extent of early coronary flow reduction in TTS and that of LV functional impairment, thus establishing some commonality of causation of the coronary and acute myocardial manifestations of TTS. However, neither extent of coronary vasculitis nor that of acute LV systolic dysfunction predict acute hypotensive changes in TTS.

**Clinical Perspective:** *What is new?:*

- We have shown for the first time, that the extent of acute coronary flow retardation in Takotsubo Syndrome (TTS) predicts that of early impairment in left ventricular (LV) systolic function.
- However, severity of acute hypotension is independent of both coronary flow reduction and LV functional impairment.

*What are the clinical implications?:*

- These results confirm that the initial pathophysiology of TTS is that of coronary vasculitis, and that the severity of this vasculitis predicts the extent of LV dysfunction.
- Therefore, future therapeutic investigations in TTS should focus on early intervention to limit coronary vasculitis.
- Hypotension/shock in acute TTS appear to have little to do with extents of acute coronary vasculitis or of LV systolic dysfunction, and therefore theoretically, neither coronary vasodilatation nor positive inotropic therapy is likely to ameliorate this problem.

## Introduction

Takotsubo Syndrome (TTS) is an acute inflammatory cardiac condition that is usually characterized by an acute coronary syndrome (ACS)-like presentation, followed by a congestive cardiac failure (CCF) aftermath(1). Like patients with acute myocardial infarction (MI), TTS patients typically present with symptoms of chest discomfort and/or dyspnea, with electrocardiographic (ECG) changes, and biochemical evidence of myocardial injury(1). There is also acute evidence of segmental (typically apical) left ventricular (LV) systolic dysfunction, which tends to resolve only partially within 3 months(2, 3), with associated symptoms of dyspnea and lethargy(4).

The major clinical problem encountered during the acute phase of TTS is symptomatic hypotension/shock, which is a marker of increased short-term mortality(5), even though it is not closely related to extent of simultaneous LV systolic dysfunction(6), nor to impairment of cardiac output(7).

Although TTS often occurs in the absence of stenoses in large epicardial coronary arteries, there is evidence of acute regional myocardial ischemia(8, 9), with variable retardation of epicardial coronary flow rates(10). These changes occur in parallel with ECG abnormalities resembling those of evolving acute MI. These findings beg the question of: What precisely causes the associated ischemia?

Pathological retardation of coronary flow rates [the “coronary slow flow phenomenon”(CSFP)(11)] may occur as a component of reperfusion injury post MI(12), but also as an infarct-independent form of acute-on-chronic small coronary artery disease(13). We have recently demonstrated that this “acute on chronic CSFP” results from microvascular spasm, with associated but reversible damage to the *endothelial glycocalyx (eGC)*, a carbohydrate-rich layer between endothelial cells and the vascular lumen(14). Damage to the eGC results in (i) impairment of vascular rheology(15) and hence theoretically increased microvascular resistance, (ii) increased expression of thioredoxin-interacting protein (TXNIP), which in turn activates the NLRP3 inflammasome(16), and (iii) increased vascular permeability(17), facilitating development of tissue edema, and extravasation of leukocytes. *All of these* have been described in the acute phases of TTS(18-20).

TTS therefore appears to present as an acute coronary “vasculitis”, and then a subsequent and prolonged myocarditis(21). If this is the case, it is possible, although currently unproven, that the extent of acute coronary vasculitis is a determinant of both severity of subsequent myocarditis (and associated LV systolic dysfunction) and of the risk of severe acute hypotension.

In the current study, utilising a large and detailed database of TTS patients, we sought to test the hypotheses that:

1. Vasculitis predicts myocarditis
  a. The extent of acute coronary flow retardation correlates directly with that of acute LV systolic dysfunction, as measured by global longitudinal strain (GLS) on echocardiography (<u>primary hypothesis</u>). Echocardiographic left ventricular ejection fraction (LVEF), a less precise marker of LV systolic function, was also assessed.
  b. The extent of acute coronary flow retardation correlates directly with other acute markers of:
    i. Biochemical evidence, and clinical and radiological consequences, of <u>eGC damage,</u> namely:
      a. Minimal systolic BP during the hospitalization
      b. Myocardial edema score on acute cardiac magnetic resonance imaging (CMR)
      c. Plasma concentration of the eGC component syndecan-1 (SD-1)
    ii. Markers of extent of myocardial injury:
      a. Peak plasma concentration of Troponin-T
      b. Peak plasma concentration of NT-proBNP
      c. Plasma concentration of the catecholamine metabolite normetanephrine
    iii. Platelet NO responsiveness, which is typically impaired proportionally to extent of redox stress(22).
  c. The extent of early coronary flow retardation <u>predicts residual impairment 3 months post TTS</u>:
    i. GLS on echocardiography
    ii. Quality of life scores as measured using the SF-36 questionnaire, with a particular focus on the physical component scores (SF36-PCS)
2. Vasculitis predicts hypotension/shock independent of myocarditis We reasoned that if the major mechanism responsible for development of acute hypotension was increased vascular permeability, this would imply that extent of hypotension might be independent of that of LV systolic dysfunction.

The results of these two types of analyses establish that (i) the extent of coronary flow retardation parallels that of acute impairment of LV systolic function in TTS patients, and (ii) that extent of acute hypotension is independent, both of impairment of LV systolic function and of coronary flow retardation.

## Methods

### Patient selection and baseline characteristics

This study was approved by the local human research ethics committee. Data from consecutive TTS patients (n=400) who presented to 3 tertiary hospitals in South Australia between May 2008 to May 2021 were assessed. Of these, patients who had early invasive coronary angiography, with adequate image quality for accurate image interpretation and frame counting were included in the analysis (n=284). A study flow chart summary is shown in Figure 1. The diagnosis of TTS was made according to the InterTAK diagnostic criteria(1). Baseline demographics, including age, sex and comorbidities including diabetes and smoking status were recorded. Data regarding predominant site of left ventricular (LV) hypokinesis and known precipitants of TTS were also recorded.

**Figure 1:**
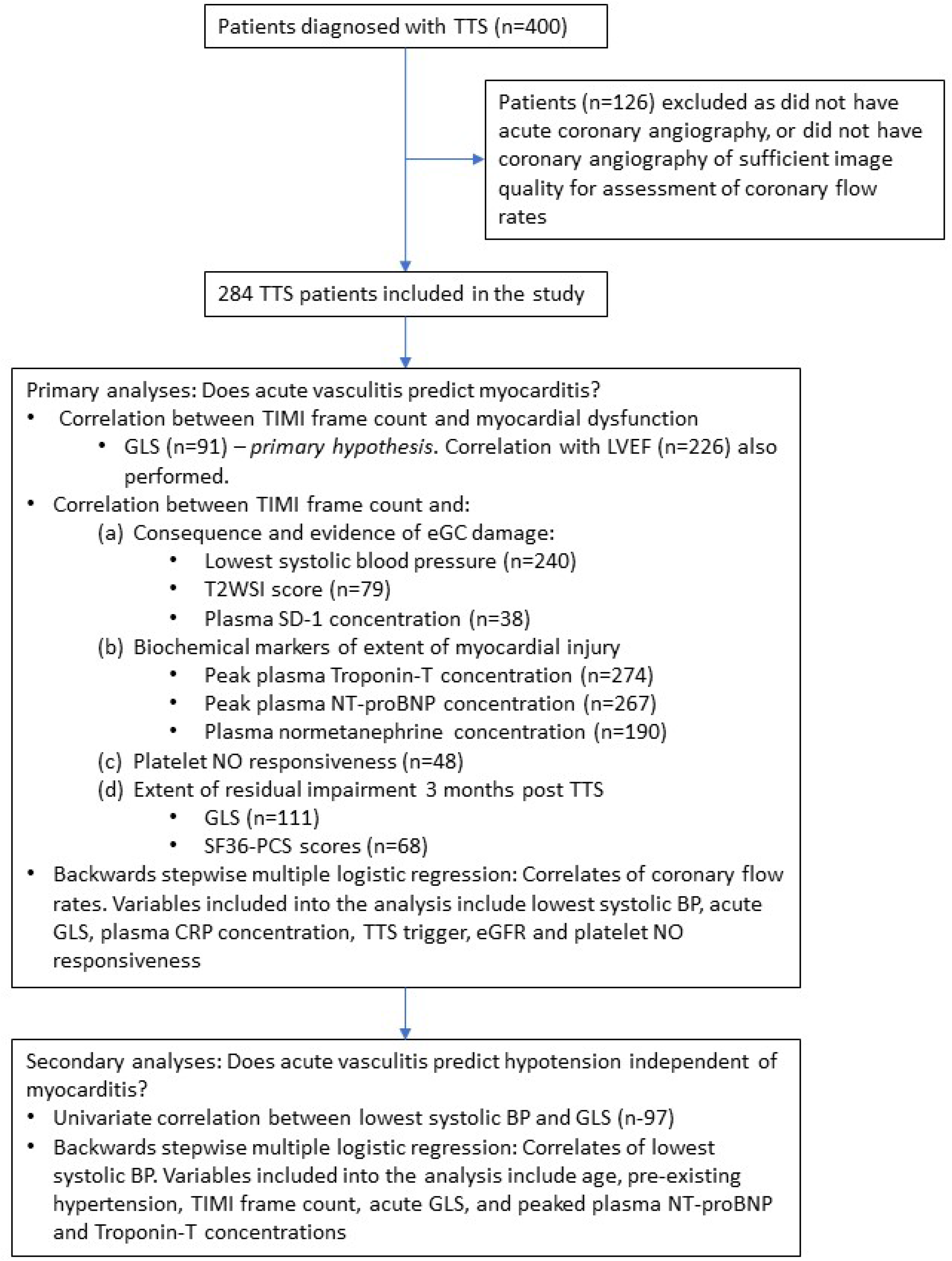
Study flow chart

Patients were routinely admitted to the coronary care or intensive care units, where blood pressures (BP) were recorded hourly, and ECGs performed at least daily. Venesection was performed routinely for measurement of hemoglobin (Hb), C-reactive protein (CRP) and estimated glomerular filtration rate (eGFR) from plasma creatinine concentrations, as well as plasma NT-proBNP, Troponin-T, and normetaphrine concentrations. In subsets of patients, the following were also measured during the first 24 hours post onset of symptoms, in all cases related to testing of specific hypotheses concerning the pathogenesis of TTS(18, 23):

1. Extent of damage to the eGC, measured using plasma SD-1 concentrations (n=38)
2. Platelet nitric oxide (NO) responsiveness, measured by the extent of inhibition of platelet aggregation with the NO donor, sodium nitroprusside (SNP) (n=48)

Plasma SD-1 concentrations were measured using immunoassay (sCD138 ELISA Kit, Diaclone Research, France). The extent of inhibition of ADP-induced platelet aggregation with SNP was measured using a dual channel impedance aggregometer (Model 560, Chrono-Log, Havertown, PA, USA), as previously described(24).

### Invasive coronary angiography

Coronary angiography was generally performed on an emergency basis. The time from hospital presentation to invasive coronary angiography was recorded. Coronary angiography was performed with standard sequences of angiographic views, and cine images were acquired at various frame rates, ranging between 10 frames per second and 30 frames per second. Coronary artery flow velocity was assessed based on TIMI Frame Count (TFC), a method previously described by Gibson et al(25). TFC is the number of cineframes required for contrast to first reach a standardized distal coronary landmark in the coronary vessel of interest. The first frame used in TFC counting is the first frame where contrast dye fully enters the coronary artery, and for the left anterior descending (LAD) coronary artery for example, the final frame is when the contrast dye reaches the most distal visible LAD bifurcation. As the length of the LAD is typically greater than the left circumflex (LCx) and the right coronary (RCA) arteries, a correction ratio of 1:1.7 is used to standardize the TFC in LAD and non-LAD vessels(25).

TFCs were measured by both FJ and GO with mean inter-observer variability of 9% of TFC. Due to the variability in image acquisition rate, the number of frame counts was adjusted to the equivalence of 30 frames per second(25).

### Echocardiography

Transthoracic echocardiography was performed in all patients at acute presentation (usually <48 hours of presentation) and again at 3 months follow-up. Standard apical two-chamber, three-chamber, and four-chamber views were obtained with special attention to LV endocardial definition, at a frame rate of at least 50 frames per second. Global longitudinal strain (GLS) was analysed using speckle-tracking software (Philips or GE) specific to each manufacturer. The endocardial border was manually traced, and region of interest was drawn to include the entire myocardium in all cases. Images were only accepted for analysis when segments approved for speckle analysis were tracked reliably. In addition, left ventricular ejection fraction (LVEF) was also measured using echocardiography, and was calculated using the biplane method.

### Cardiac magnetic resonance (CMR) imaging

CMR was performed in patients without any contraindications to this procedure during their initial hospital admission, on a 1.5T Philips MRI system, with a five-channel phased-array coil and electrocardiographic gating. T2-weighted-Black-Blood images were analysed to determine the extent of myocardial inflammation, utilising certified MRI evaluation software [OsiriX Lite (http://www.osirix-viewer.com/)]. In each patient, short axis views of the left ventricle were obtained at the apex, mid and base, and the endocardial and epicardial contours were traced manually in all 3 levels. In order to provide a measure of total LV edema, we utilized methodology previously developed by Neil et al(26): Mean signal intensity (SI) score was determined for each short-axis slice, and results were expressed relative to the SI score of their spleen, giving a SI ratio (or a “T2WSI score”).

### Follow up

Patients were followed up 3 months after the acute TTS attack, with LV systolic function measured by GLS, and quality of life assessed using the SF-36 Questionnaire with a particular focus on the physical component scores (SF36-PCS), both included in the analysis.

### Statistical analyses

To test the <u>central hypothesis</u> that extent of coronary microvascular dysfunction during acute TTS is directly associated with the extent of myocardial dysfunction, correlations were sought between TFC on invasive coronary angiography with GLS on echocardiography. Correlation analyses were performed by Pearsons’s or Spearman’s correlation coefficients as appropriate. Correlations were also sought for the secondary measures mentioned above and listed in Figure 1. Associations between TFC and patient demographics were also sought with unpaired t-test and Wilcoxon-Mann-Whitney test as appropriate. Subsequently, those univariate parameters with a p-value ≤0.10 were included in a multivariate backwards stepwise multiple logistic regression analysis to identify multivariate correlates of TFC.

In order to test the subsidiary hypothesis that extent of acute hypotension is independent of extent of impairment in GLS, we performed univariate correlations of minimal systolic BP with GLS, followed by backwards stepwise multiple logistic regression analysis, utilizing minimal systolic BP as the dependent variable, and including age, pre-existing diagnosis of hypertension, average (LAD-corrected) TFC, acute GLS, and peaked plasma NT-proBNP and Troponin-T concentrations as potential correlates.

Data are presented as mean ± SD or median (inter-quartile range) as appropriate, and the limit of statistical significance was set at p<0.05.

## Results

### Patient characteristics and coronary angiography findings

A total of 284 TTS patients were included in the analysis. Patient demographics are summarized in Table 1. As expected, the majority of patients were older females, with few conventional cardiovascular risk factors other than advanced age.

**Table 1:** Patient demographics. Non-Gausian data are shown as median (IQR).

|  |  |
| --- | --- |
|  | Total patients (n=284) |
| Age (years) | 68 (60-76) |
| Female, n (%) | 268 (94%) |
| TTS triggered by a life-threatening physical illness, n (%) | 68 (24%) |
| Apical TTS, n (%) | 178 (63%) |
| Diabetes Mellitus, n (%) | 41 (14%) |
| Hypertension, n (%) | 135 (48%) |
| Current smoker, n (%) | 36 (13%) |
| Hemoglobin, g/L | 134 (126-142) |
| Plasma CRP, mg/L | 9.5 (3.6-31) |
| Estimated GFR, mL/min | 67 (60-90) |
| In-hospital mortality, n (%) | 4 (1.4%) |

153 (53%) patients had an invasive coronary angiogram within the first 24 hours of their presentation. Frame counts did not differ significantly between patients who had angiography within 24 hours and those who did not.

The corrected TFC of the LAD correlated closely with the averaged TFC in other major epicardial arteries (r=0.83, p<0.0001). Given the fact that TTS-associated myocardial edema is seen not just in the hypokinetic left ventricular segments, but also throughout the myocardium(26), the averaged (LAD-corrected) TFC of all coronary arteries was chosen as the main dependent variable.

Median averaged (LAD-corrected) TFC of all 3 major coronary arteries was 33 frames (IQR 23.5-37.7), which is higher compared to previously reported data for healthy subjects (<21 frames)(25).

TFC did not significantly correlate with age (p=0.23), gender (p=0.20), smoking status (p=0.9) or diabetes status (p=0.50). TFC also did not differ between apical and non-apical forms of TTS (p=0.21). However, TFC was less elevated (median 30 vs 33 frames, p=0.003) in patients with TTS precipitated by a physical trigger than in other subgroups (p=0.03).

Renal function was preserved in the majority of patients (median eGFR 67mL/min), and TFC varied directly with eGFR on univariate correlation (r=0.24, p<0.0001)(Figure 2A). Plasma CRP concentration correlated inversely with TFC (r=-0.21, p=0.001).

**Figure 2:**
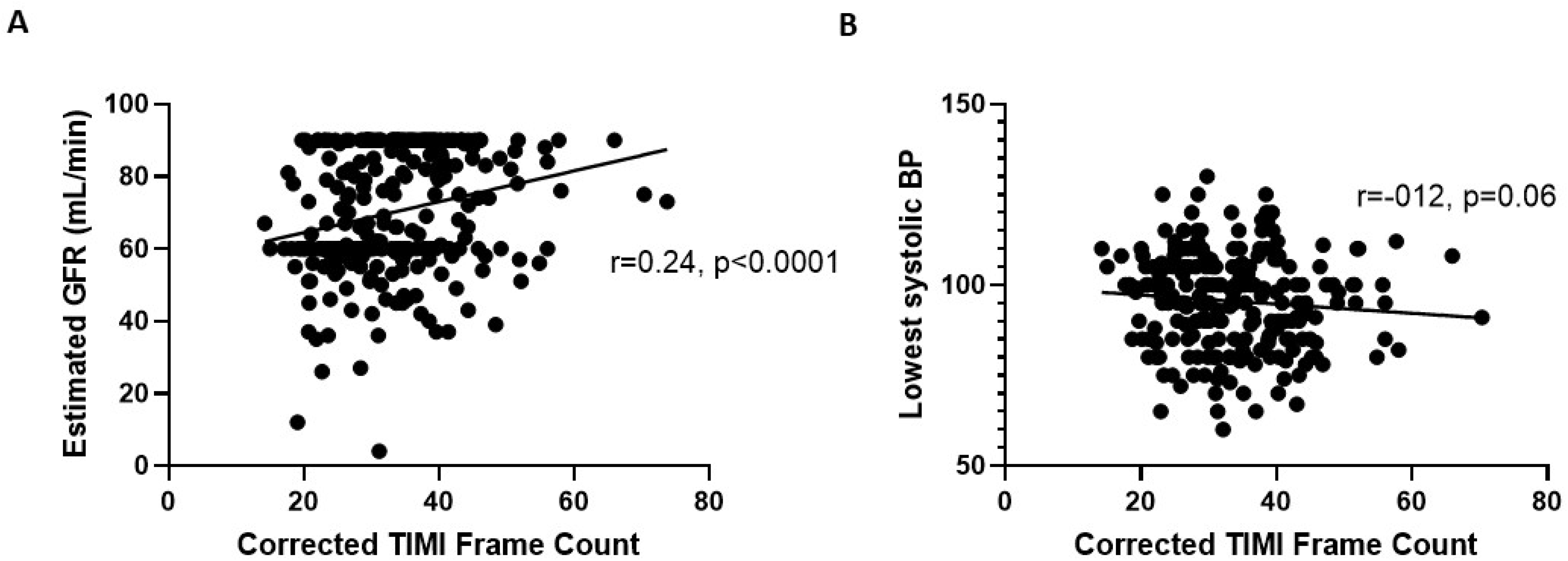
Association between corrected TIMI frame counts and A) estimated glomerular filtration rate (eGFR), and B) lowest systolic blood pressure. Laboratory methodology includes truncation of eGFR values at maximum of 90mL/min.

### Primary hypothesis: Does acute vasculitis predict subsequent myocarditis?

#### (a) CSFP predicts extent of acute systolic functional impairment

The extent of coronary flow retardation correlated directly with that impairment of LV systolic function on index echocardiography, as measured by GLS on univariate analysis (Figure 3). However, this trend did not approach statistical significance when LVEF replaced GLS (p=0.27).

**Figure 3:**
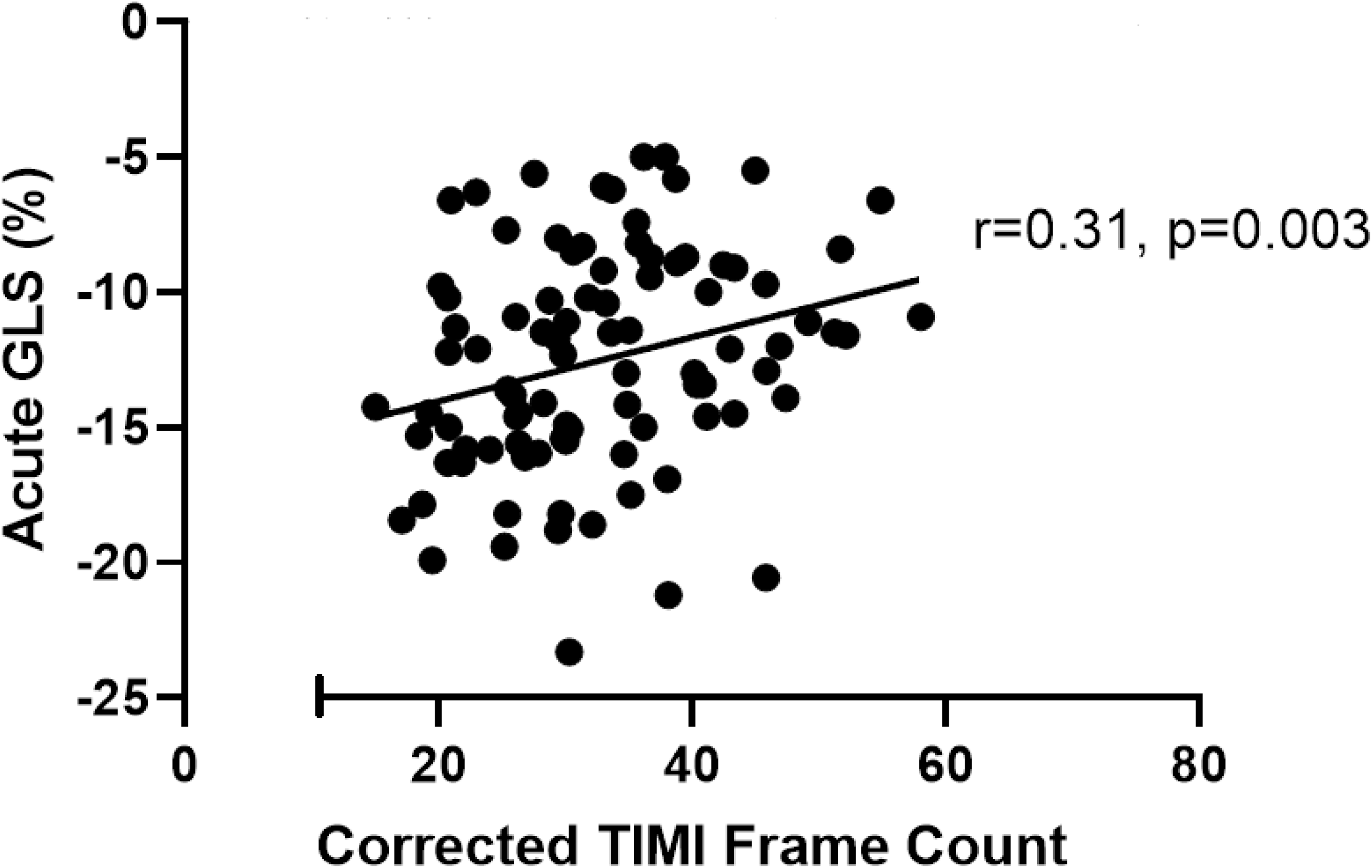
Primary analysis: association between corrected TIMI frame counts and acute left ventricular global longitudinal strain (GLS).

#### (b) Other acute comparisons

Hypotension (systolic BP ≤90mmHg) occurred in 37% of patients, with mean nadir of systolic BP of 96mmHg in the studied population. Patients who had lower systolic BP tended to have higher TFC (r=-0.12, p=0.06) (Figure 2B). There was however no significant correlation between TFC and either T2WSI score on CMR (p=0.75) or plasma SD-1 concentration (p=0.33).

Biochemical markers of extent of acute myocardial injury (peak plasma Troponin-T, NT-proBNP, and normetanephrine concentrations) also did not correlate significantly with TFC (p=0.43, p=0.16, and p=0.37 respectively).

In a subset of patients (n=48) where platelet NO responsiveness was measured, whilst there was no statistically significant correlation between TFC and NO responsiveness, there was a trend towards greater flow retardation(p=0.07) in patients with most pronounced responsiveness to NO.

#### (c) Indices of late recovery

At 3 months’ follow up, mean GLS was -17.91% ± 3.14%, consistent with persistent but mild LV systolic dysfunction in many patients. Similarly, quality of life scores were also mildly impaired, with a mean SF36-PCS score of 56.1, as compared to the “normal” mean value of 65 for the female population aged 65-74 of the region of sampling(27). However, there was no statistically significant correlation between extent of acute coronary flow retardation and either GLS or quality of life scores at 3 months post-TTS (p=0.60 and p=0.67 respectively).

#### (d) Multivariate analysis

Acute GLS, TTS triggers, eGFR, lowest in-hospital systolic BP, and plasma CRP concentration were included in a backwards stepwise multiple logistic regression analysis, where the dependent variable was corrected TFC. The result of this multivariate analysis is summarized in Table 3. Both acute GLS and eGFR were independent and direct correlates of TFC acutely (β=0.52, p=0.03 and β=0.50, p=0.02 respectively). Surprisingly there was as trend (p=0.06) for low CRP values to correspond to high acute TFC. Importantly, there was no correlation between minimal systolic BP and TFC.

**Table 2:** Acute correlates of frame count: final iteration of backwards stepwise multiple logistic regression analysis.

| Variable | $\beta$ | p-value |
| --- | --- | --- |
| Acute GLS | 0.52 | 0.03 |
| Plasma CRP concentration | -0.44 | 0.06 |
| Estimated GFR | 0.50 | 0.02 |

### Secondary hypothesis: Vasculitis predicts hypotension/shock independent of myocarditis

The issue was approached by initial univariate correlation between TFC and minimal systolic BP followed by multivariate analysis of correlates of minimal systolic BP. As previously mentioned, TFC tended (r=-0.12, p=0.06) to be inversely correlated with minimal systolic BP (Figure 2B). However, there was no significant univariate correlation between GLS and minimal systolic BP (data not shown). On multivariate analysis, the absence of previous systolic hypertension, and extent of acute Troponin-T rise were the only identified significant correlates of minimal systolic BP (β=0.33, p=0.002 and β=-0.27, p=0.01 respectively).

## Discussion

The precise pathophysiology of what is now usually named “Takotsubo Syndrome” remains uncertain. The Japanese investigators who initially described the condition, including the eponymous distortion of ventricular contraction, considered the ACS-like presentation and suggested that the pathogenesis might be of multivessel coronary spasm(28). In retrospect, this was surprising, because coronary artery spasm occurs very commonly in Japan(29), and does not usually lead to myocardial necrosis(30). Conversely, a subsequent generation of (largely European) investigators focused on the emergence, typically during the late hospitalization period, of heart failure, and categorized the condition as a congestive cardiomyopathy (TTC), a term which only recently has declined in use(31).

Very recently, we have proposed that *TTS begins as an acute coronary vasculitis, and evolves into a myocarditis of variable severity and duration*(21). However, little has been done to determine whether the putative second phase results from the first. In the current study, our primary hypothesis was that extent of coronary flow retardation would predict that of early GLS impairment. This proved to be the case. Thus, the rapid development of segmental LV systolic dysfunction either directly reflects an identical stimulus, and/or represents a predictable consequence of early coronary vasculitis.

Whilst injury to the vascular eGC may have contributed to impaired myocardial contractility via vascular permeabilization(17), for example by facilitating monocyte and macrophage transmigration(20), and increasing expression of the humoral pro-inflammatory protein TXNIP(16), it is equally probable that flow retardation per se may have attenuated the Gregg phenomenon(32), and thus impaired LV contractility.

Subsidiary investigations undertaken in this context sought correlations between TFC and a number of biochemical parameters. The only significant correlations found were with CRP concentrations, which were inversely related to TFC, and with eGFR, which was strongly and directly related to TFC. The reasons underlying those findings are not clear-cut. As regards eGFR, a possible explanation for this finding may be that hemodynamic changes in patients with CKD induce physiological adjustments similar to those occurring in the “diving reflex”, in that peripheral (catecholamine-induced) vasoconstriction predominates in patients with CKD, leading to redistribution of blood flow to adequately perfuse the heart and the brain(33). Indeed, there is also evidence of increased resting cerebral blood flow in patients with CKD(34). The lack of an explanation for the findings regarding CRP correlations may relate to the precipitation of TTS in a minority of patients by infections, but this remains speculative at this stage. More extensive TFC prolongation tended to be associated (p=0.07) with platelet hyper-responsiveness to NO, which has been implicated as being generated via aberrant β-adrenoceptor stimulation and NO synthase activation(35, 36). This forms a basis of peroxynitrite generation and thus incremental oxidative stress(19). This marginal association is therefore not surprising.

Additionally, there was no correlation between extent of acute flow retardation and that of LV systolic dysfunction and quality of life impairment after 3 months. Data from Dawson’s group have revealed that both persistent myocardial energetic impairment and the emergence of myocardial fibrosis are prominent late pathogenetic features in TTS(3): presumably neither of these are closely related to extent of acute coronary vasculitis.

A secondary purpose of the current study was to provide additional understanding of the factors responsible for the development of severe hypotension in the early stages of TTS. We have previously shown that occurrence of severe hypotension is independent of acute changes in pulmonary artery oxygen saturation (a marker of cardiac output)(7) and is statistically related to fall in LVEF, but only weakly so(6). The current analysis showed that minimal systolic blood pressure is significantly lower in patients without past histories of hypertension, and also in patients with greater increases in plasma troponin-T concentrations. The latter data are not unexpected, and unfortunately the hypotension:troponin release nexus does not help elucidate cause vs effect. The current quantitative and multivariate analysis does not provide significant clarification regarding mechanism(s) of hypotension, which was independent both of TFC and of GLS. Unfortunately, the current methodology did not permit us to measure extent of fluid extravasation associated with coronary microvascular injury: this might have provided significant mechanistic information. Nevertheless, the current data provide additional evidence that extent of hypotension is not closely related to that of LV systolic dysfunction.

The issue of coronary vasculitis as a basis for subsequent myocardial dysfunction in TTS has been appreciated in two previous studies, independent of our previous postulate of sequential pathogenesis(21). In 2020, Montone et al. demonstrated in a cohort of 101 TTS patients, that acute coronary flow retardation (TIMI-2 flow) was associated with an increased risk of in-hospital acute heart failure(37). Sans-Rosello et al. evaluated changes in coronary microvascular resistance in 166 TTS patients, and showed that increased microvascular resistance predicted the occurrence of symptomatic heart failure (usually mild) over subsequent 12 months(38). The importance of this study can now be seen as providing an extension of our current acute/subacute findings for a period of at least 12 months(39). Finally, a very recent report(40) in a mouse model of transaortic constriction demonstrated gradual development of LV apical hypokinesis, which was reversed by 2-4 weeks’ treatment with the coronary vasodilator agent chromonar. Although it may be hazardous to extrapolate from this experimental design to TTS, the findings might be interpreted as additional evidence of a nexus between reversal of early coronary vasoconstriction and of impaired systolic function in a “TTS-like” model.

There are several caveats to our study. Firstly, due to the retrospective design of our study, our finding of directly related impairment of coronary flow and myocardial dysfunction implies only association, rather than causation. It might be possible to test Koch’s postulates regarding causation by evaluating the impact of eGC-protecting agents, such as the matrix metalloproteinase inhibitor doxycycline(41), the hydrogen sulphide donor N-acetylcysteine(42), or even potentially sodium-glucose cotransporter-2 inhibitors (SGLT-2i) such as empagliflozin(43).

Secondly, flow retardation as measured by TFC does not always represent microvascular dysfunction, and other potential contributors such as coronary vascular ectasia(44), or extramural compression of coronaries in association with diastolic dysfunction(45) may need to be considered. Furthermore, myocardial edema quantitation utilized T2-weighted signal intensity score measurement, as in our previous report(26):- it is possible that more recently developed techniques such as T2-mapping sequences might have been more precise(46). Finally, as the data on some secondary end points such as plasma SD-1 concentrations, myocardial edema score on CMR, and platelet NO responsiveness were only available in small numbers of patients, these analyses were subject to Type 2 error.

In conclusion, we have shown for the first time, an acute association between coronary flow retardation and myocardial systolic dysfunction in patients with TTS. Furthermore, the extent of flow retardation is not a significant correlate of severity of hypotension. Thus, our results establish a nexus between coronary vasculitis and myocarditis as a central component of the pathogenesis of TTS, and suggest that limitation of coronary microvascular dysfunction may represent an important early therapeutic target for patients with TTS in the future.

## Data Availability

The data that support the findings of this study are available from the corresponding author upon reasonable request.

## Acknowledgements

We wish to acknowledge the contributions of Dr Yuliy Chirkov and Dr Ha Nguyen, who performed the investigations of platelet physiology and of plasma SD-1 concentrations. We also are grateful to the medical and nursing staff, as well as echocardiography and CMR technicians at Lyell McEwin Hospital, Queen Elizabeth Hospital, and Royal Adelaide Hospitals. Finally, we are very grateful to Ms Jeanette Stansborough for her substantial contributions to patient recruitment and data entry.

